# Racial disparities in prevalence, determinants, and impacts of COVID-19 in pregnancy: Protocol for a study using data from New Jersey hospitals

**DOI:** 10.1101/2021.05.23.21257672

**Authors:** Slawa Rokicki, Pauline Nguyen, Alaine Sharpe, Dyese Taylor, Suzanne Spernal, Archana Raghunath, Leslie Kantor

**Author notes:** **Corresponding author: Slawa Rokicki**, Rutgers School of Public Health, 683 Hoes Lane West, Office 232, Piscataway, NJ 08854.

## Abstract

**Introduction:** Racial and ethnic disparities in COVID-19 related infections, hospitalizations, and deaths have been well-documented. However, little research has examined racial and ethnic disparities in COVID-19 prevalence, determinants, and impacts among pregnant women. Within the United States, New Jersey was an early epicenter of the pandemic and experienced high rates of disease in the fall of 2020.

**Methods:** This study uses data from two New Jersey hospitals, which implemented universal testing of COVID-19 of pregnant women admitted for labor and delivery starting in March 2020. We will estimate prevalence of COVID-19 between March 2020 and November 2020 and compare prevalence rates across race and ethnicity. We will conduct multivariable logistic regression analysis to examine the associations of COVID-19 infection with patient demographic and health status predictors. We will also use multivariable linear and logistic regressions to examine the impact of COVID-19 symptomatic and asymptomatic infection on maternal and infant birth outcomes.

**Discussion:** This study will generate important policy implications on birth equity in the time of COVID-19 and guide future research studies related to COVID-19 in pregnant women. Results of this study will help to guide interventions and policies to center safe, accessible, and equitable maternity care within the strategic response to the pandemic.

## Introduction

Since its emergence in late 2019, the severe acute respiratory syndrome coronavirus 2 (SARS-CoV-2) that causes novel coronavirus disease (COVID-19) has infected over 163,720,868 people worldwide, approximately one-fifth of whom are in the United States (32,972,982) as of early May 2021.^1–4^ Within the United States, New Jersey was an early epicenter of the pandemic, and once again experienced high rates of disease in the fall of 2020.^5^

Despite significant advancements in our understanding of SARS-CoV-2 infection, the effects of COVID-19 on pregnant women and pregnancy outcomes remains understudied. Due to altered physiology and immune function, pregnant women and their fetuses are generally considered a high-risk population during infectious disease outbreaks.^6–10^ Available evidence suggests that the incidence of preterm births, low birth weight, Cesarean section, and ICU admission appears higher among pregnant women with COVID-19 than among women without COVID-19.^8,11–16^

Racial disparities in COVID-19 related infections, hospitalizations, and deaths have been well-documented.^17,18^ Disparities reflect the health consequences of the social, environmental, and structural effects of racism, including differences in prevalence of underlying chronic conditions and exposure to impacts of the social determinants of health including access to medical insurance, wealth, and employment as essential employees.^19,20^ However, few studies have examined racial disparities in perinatal outcomes of COVID-19.^19,21^ More research is critical for developing clinical care guidelines that promote birth and reproductive health equity.

New Jersey is a particularly important setting to study disparities in COVID-19 during pregnancy because of its diverse population and the high incidence of COVID-19 that affected the population of New Jersey in the early months of the pandemic.^5^ To date, there have been few studies on COVID-19 in pregnant women in New Jersey.^22^ Little is known about how many pregnant women in New Jersey contracted the disease at various time points in the pandemic, the factors that affect likelihood of infection in pregnant women, and whether symptomatic status and pre-existing conditions influence the outcomes of their pregnancies or their infants’ health. This study will examine these research questions using data from hospital records in a region with high incidence of COVID-19 that implemented universal COVID-19 testing of pregnant women.

## Methods

### Goal and objectives

The study will use data from two New Jersey hospitals collected between March and November 2020. The primary study objectives are:

1. To determine the prevalence of COVID-19 in pregnant women in two New Jersey hospitals and assess how prevalence changed from March to November 2020
2. To assess disparities in rates by race and ethnicity, geography of residence (ZIP code), age, body mass index (BMI), length of gestation, parity, insurance status, and pre-existing health conditions
3. To examine differences in maternal and infant health outcomes among women positive for COVID-19 who were symptomatic on admission, women positive for COVID-19 who were asymptomatic on admission, and women negative for COVID-19

Secondary study objectives are:

1. To examine differences in maternal and infant health outcomes between diabetic pregnant women with symptomatic and asymptomatic COVID-19 and diabetic women negative for COVID-19

### Study population

The study will use data from the electronic medical records (EMR) of all women who were admitted for labor and delivery experience at the Newark Beth Israel (NBI) and Clara Maass Medical Centers (CMMC), between March 17th, 2020 and November 20th, 2020. Both hospitals are located in Essex county, New Jersey, which has a diverse and low-income population. In 2019, 21% of births in Essex county were to White non-Hispanic mothers, 40% were to Black non-Hispanic, 30% were to Hispanic mothers of any race, and 9% were to Asian or other races. ^23^ 41% of all births were to mothers with Medicaid insurance. 46% of births were to foreign-born mothers.

### Project management

Our collaborative multi-site research team brings together the Rutgers University School of Public Health, NBI, and CMMC. The partners from these hospitals are involved in the research and will collaborate on the study design, data extraction and transfer, and interpretation of results.

### Data and measures

Both NBI and CMMC implemented universal testing of COVID-19 of pregnant women admitted for labor and delivery in March 2020. Data will be extracted from two sources: (1) NBI electronic medical records and (2) CMMC electronic medical records. The medical records reflect birth hospitalizations, including both women’s records and infant records. Hospital administrators in each hospital will provide access to these records. The records will be extracted by our partners on the research team at each respective hospital and transferred to the Rutgers School of Public Health. We anticipate 1300 records from each hospital, with approximately 50 COVID positive cases.

The following variables will be extracted from the electronic medical record. COVID-19 status will be extracted and categorized as positive symptomatic, positive asymptomatic, negative. Demographic and health status variables extracted include date of admission, race/ethnicity, ZIP code, age (in years), BMI, length of gestation upon admission for labor and delivery, parity, insurance status, birth weight, smoking status, history of preterm birth, history of hypertension, and history of diabetes. Outcome variables include type of delivery (vaginal, c-section), depression/anxiety (as measured by the Postnatal Edinburgh Depression Scale^24^), admittance to ICU (mother), any supplemental oxygen used, vascular disease, fetal distress, newborn respiratory distress syndrome, admission to NICU (infant), APGAR score (infant), breastfeeding status, and infant weight gain.

### Ethics approvals and data security considerations

Institutional Review Board (IRB) approvals have been obtained at each institution (Rutgers School of Public Health ID# Pro2020002959; NBI IRB#2021.19; CMMC IRB R2021-01CMMC). As this study is a retrospective chart review, an exemption for consent to access patient data was obtained. This research study presents minimal risks to study subjects. We will not collect subject names, social security numbers, or medical record numbers but the data will include protected health information in the form of ZIP code and admission date. The shared file will contain random identifier numbers and will be password-protected. The data will be kept securely for 6 years after study closure.

### Data analysis plan

The data analysis will include descriptive statistics (means, medians, standard deviations) of all predictor and outcome variables. We will compare prevalence of COVID-19 over time and across race/ethnicity groups, and compare the prevalence to that of the general population using publicly available data from New Jersey Department of Health.^5^ We will conduct multivariable logistic regression analysis to examine the associations of COVID-19 infection with patient demographic and health status predictors. We will also explore use of multinomial logistic regression to examine determinants across the three categories of COVID-19 infection. Finally, we will use multivariable linear (continuous outcomes) and logistic (binary outcomes) regression to examine the impact of COVID-19 infection (as categorized by symptomatic, asymptomatic, or negative) on maternal and infant birth outcomes. Data analysis will be conducted using Stata v16.

### Dissemination of results

Results will be disseminated in peer reviewed scientific journals and at academic conferences. Research findings will be communicated to relevant state and local policy makers.

### Discussion

This multi-site collaborative research study will examine racial disparities in the prevalence of COVID-19 in pregnant women and examine determinants and outcomes of COVID-19 in pregnant women and infants using secondary data from birth records at two New Jersey hospitals with diverse and low-income populations. This study is one of the first to examine SARS-CoV-2 infection prevalence and outcomes among pregnant women in New Jersey, a state that experienced high rates of infection early in the pandemic.

There is a growing body of evidence that COVID-19 has significant health consequences for pregnant women and infants, with increases in maternal deaths, stillbirth, ruptured ectopic pregnancies, and maternal depression.^6,15,16^ However, little research has examined racial/ethnic disparities in COVID-19 prevalence, determinants, and impacts in pregnancy.^19,21^

This study will generate important policy implications on birth equity in the time of COVID-19 and guide future research studies related to SARS-CoV-2 infection in pregnant women. The study outcomes will help to illuminate larger community prevalence rates and provide insights on population disparities and equity issues among pregnant women. Results of this study will help to guide interventions and policies to center safe, accessible, and equitable maternity care within the strategic response to the pandemic, including the allocation of health care resources to heavily affected populations.

## Data Availability

Data is not available to the public as data are obtained electronic medical records and a data use agreement with the hospitals involved is necessary to use the data.

## Notes

### Competing Interest Statement

The authors have declared no competing interest.

### Funding Statement

No external funding was received.

### Author Declarations

Institutional Review Board (IRB) approvals have been obtained at Rutgers School of Public Health ID# Pro2020002959; Newark Beth Israel Medical Center IRB#2021.19; Clara Maass Medical Center IRB R2021-01CMMC.

